# Clinical Characteristics and Short-Term Outcomes of Severe Patients with COVID-19 in Wuhan, China

**DOI:** 10.1101/2020.04.24.20078063

**Authors:** Xiaobo Feng, Peiyun Li, Liang Ma, Hang Liang, Jie Lei, Wenqiang Li, Kun Wang, Yu Song, Shuai Li, Wei Yang, Cao Yang

## Abstract

**Objective:** A novel pneumonia (COVID-19) which is sweeping the globe was started in December, 2019, in Wuhan, China. Most deaths occurred in severe and critically cases, but information on prognostic risk factors for severe ill patients is incomplete. Further research is urgently needed to guide clinicians, so we prospectively evaluate the clinical outcomes of 114 severe ill patients with COVID-19 for short-term in the Union Hospital in Wuhan, China.

**Methods:** In this single-centered, prospective and observational study, we enrolled 114 severe ill patients with confirmed COVID-19 from Jan 23, 2020 to February 22, 2020. Epidemiological, demographic and laboratory information were collected at baseline, data on treatment and outcome were collected until the day of death or discharge or for the first 28 days after severe ill diagnosis, whichever was shorter. Univariate and multivariate Cox proportional hazard models were used to determine hazard ratios (HRs) and 95% confidence intervals (CIs) of poor outcome.

**Results:** Among enrolled 114 patients, 94 (82.5%) had good outcome while 20 (17.5%) had poor outcome. No significant differences were showed in age, gender and the prevalence of coexisting disorders between outcome groups. Results of multivariate Cox analyses indicated that higher levels of oxygen saturation (HR, 0.123; 95% CI, 0.041-0.369), albumin (HR, 0.060; 95% CI, 0.008-0.460) and arterial partial pressure of oxygen (HR, 0.321; 95% CI, 0.106-0.973) were associated with decreased risk of developing poor outcome within 28 days. In the other hand, higher levels of leucocytes (HR, 5.575; 95% CI, 2.080-14.943), neutrophils (HR, 2.566; 95% CI, 1.022-6.443), total bilirubin (HR, 6.171; 95% CI, 2.458- 15.496), globulin (HR, 2.526; 95% CI, 1.027-6.211), blood urea nitrogen (HR, 5.640; 95% CI, 2.193-14.509), creatine kinase-MB (HR, 3.032; 95% CI, 1.203-7.644), lactate dehydrogenase (HR, 4.607; 95% CI, 1.057-20.090), hypersensitive cardiac troponin I (HR, 5.023; 95% CI, 1.921-13.136), lactate concentration (HR,15.721; 95% CI, 2.099-117.777), Interleukin-10 (HR, 3.551; 95% CI, 1.280-9.857) and C-reactive protein (HR, 5.275; 95% CI, 1.517-18.344) were associated with increased risk of poor outcome development. We also found that traditional Chinese medicine can significantly improve the patient’s condition, which is conducive to the transformation from severe to mild.

**Conclusion:** In summary, we firstly reported this single-centered, prospective and observational study for short-term outcome in severe patients with COVID-19. We found that cytokine storm and uncontrolled inflammation responses, liver, kidney, cardiac dysfunction may play important roles in final outcome of severe ill patients with COVID-19. Our study will provide clinicians to be benefit to rapidly estimate the likelihood risk of short-term poor outcome for severe patients.

## Introduction

An outbreak of pneumonia caused by a novel coronavirus, severe acute respiratory syndrome corona virus 2 (SARS-CoV-2), centered in Wuhan, Hubei province, in December 2019.[1] It has rapidly spread to other cities and countries outside Wuhan and has become a global health crisis. As the disease progresses, more and more characteristics about the disease are known to us. Up to now, there are still no specific drugs, mainly supporting treatment in clinic. A large number of studies have shown that the main symptom of this disease was fever, cough and dyspnea.[2-5] Some articles have described the disease in details. Huang et al described epidemiological, clinical, laboratory, and radiological characteristics, treatment, and outcomes of 41 patients that were first group admitted hospital patients and diagnosed with SARS-CoV-2 infection, and they also compared the clinical characteristic between intensive care unit (ICU) and non-ICU patients.[2] Yang et al made a detailed analysis about the patients with critically ill with SARS-CoV-2 infection.[5] Jin et al descripted the characteristics of 74 cases of coronavirus infected disease 2019 (COVID-19) with gastrointestinal symptoms and proposed that non-classical symptoms have been overlooked, posing a threat to the public.[6] Wu et al discussed that older people (⩾65 years old) with greater risk of development of acute respiratory distress syndrome (ARDS) and death.[7] Guo et al found that diabetes is a risk factor for the patients with COVID-19.[8] However, few studies took prospective study to explore the short-term outcomes of severe ill patients under current medical treatment and the risk factors that affect the short-term outcomes of severe ill patients, especially pneumonia patients with certain chronic diseases, which accounted for the majority of deaths. Here, we used a single-centered, prospective method to describe the basic clinical characteristics and short-term outcomes of severe patients in Union hospital, Wuhan, and further explore the potential risk factors for poor outcome through Cox proportional hazard models.

## Methods

### Study design and participants

This is a single-center, prospective study of 114 severe patients with confirmed COVID-19 pneumonia hospitalized at Union Hospital in Wuhan, China, which is a designated hospital to treat patients with COVID-19. We continuously enrolled patients from January 23, 2020 to February 22, 2020, who had been diagnosed with COVID-19, according to WHO interim guidance. Based on the Diagnosis and Treatment Scheme for SARS- CoV-2 of Chinese (The Seven Edition), severe patients were diagnosed if one or more following criteria were met: dyspnoea with respiratory rate (RR) ≥ 30 times/min, resting finger oxygen saturation ≤ 93%, artery PaO 2 /FiO 2 ≤ 300 mm Hg (1 mm Hg=0.133 kPa). This study was approved by the Ethics Commission of Wuhan Union Hospital of Tongji Medical College, Huazhong University of Science and Technology. Written informed consent was waived for the emergency of this infectious diseases.

### Data collection

Data on clinical characteristics were collected by using a case record form modified from the standardized International Severe Acute Respiratory and Emerging Infection Consortium case report forms. Epidemiological and demographic information including age, sex, coexisting disorders were also collected. The baseline laboratory indices and radiographic results were obtained from clinical electronic medical records. Moreover, the treatment and outcome data were collected until the day of death or discharge or for the first 28 days after severe ill diagnosis, whichever was shorter. All missing or vague data, direct communications with patients and their families. All data were finally checked by two physicians (Xiaobo Feng and Liang Ma) and a third researcher (Wei Yang) adjudicated any difference in interpretation between the two primary reviewers.

### Outcomes

Clinical outcomes of 28 days’ consecutive observations of severe patients were divided into two categories, including good outcome (discharge, non-severe or ventilator free) and poor outcome (mechanically ventilated or dead). The criteria of discharge including the temperature returned to normal for more than 3 days (T < 37.3°C), respiratory symptoms were improved significantly, pulmonary imaging showed significant improvement in acute exudative lesions and nucleic acid test was negative for respiratory tract specimens such as sputum and nasopharyngeal swab for two consecutive times (at least 24 hours after sampling). Non-severe mean mild type which is defined as having slight clinical symptoms without pneumonia on radiography.

### Statistics

Continuous variables were expressed as means ± SDs if normally distributed and medians (IQRs) if skewed distributed while categorical variables were summarized as number (%). Differences of characteristics between outcome groups were assessed using student’s *t* test or Mann-Whitney U test for continuous variables, and chi-square test for categorical variables.

In addition, univariate and multivariate Cox proportional hazard models were used to determine hazard ratios (HRs) and 95% confidence intervals (CIs) of poor outcome in severe patients with COVID-19. The candidate risk factors included demographic and epidemiological characteristics, as well as some laboratory indices. We determined the cut points of levels according to normal range, actual distribution and clinical significance of each index. Adjustment were made for potential confounders including age and sex. For risk factors identified in Cox analyses, we used restricted cubic spline model to further explore the potential dose-response relationship between factors and poor outcome risk. The referent (HR = 1) was set according to the cut point in Cox analyses. *P* < 0.05 was considered statistically significant. All data were analyzed by SPSS (23.0 IBM SPSS).

## Results

### Clinical Outcomes

A total of 114 patients with COVID-19 were enrolled in this study. At 28 days after severe ill diagnosis, 94 patients were considered as had good outcome while 20 patients had poor outcome. As shown in Figure 1, 51 (45%) patients were alive and discharge, 39 (34%) turned to non-severe ill, 4 (3%) remained severe status and ventilator free, 11 (10%) were alive but still ventilated and 9 (8%) patients had died.

**Figure 1.**
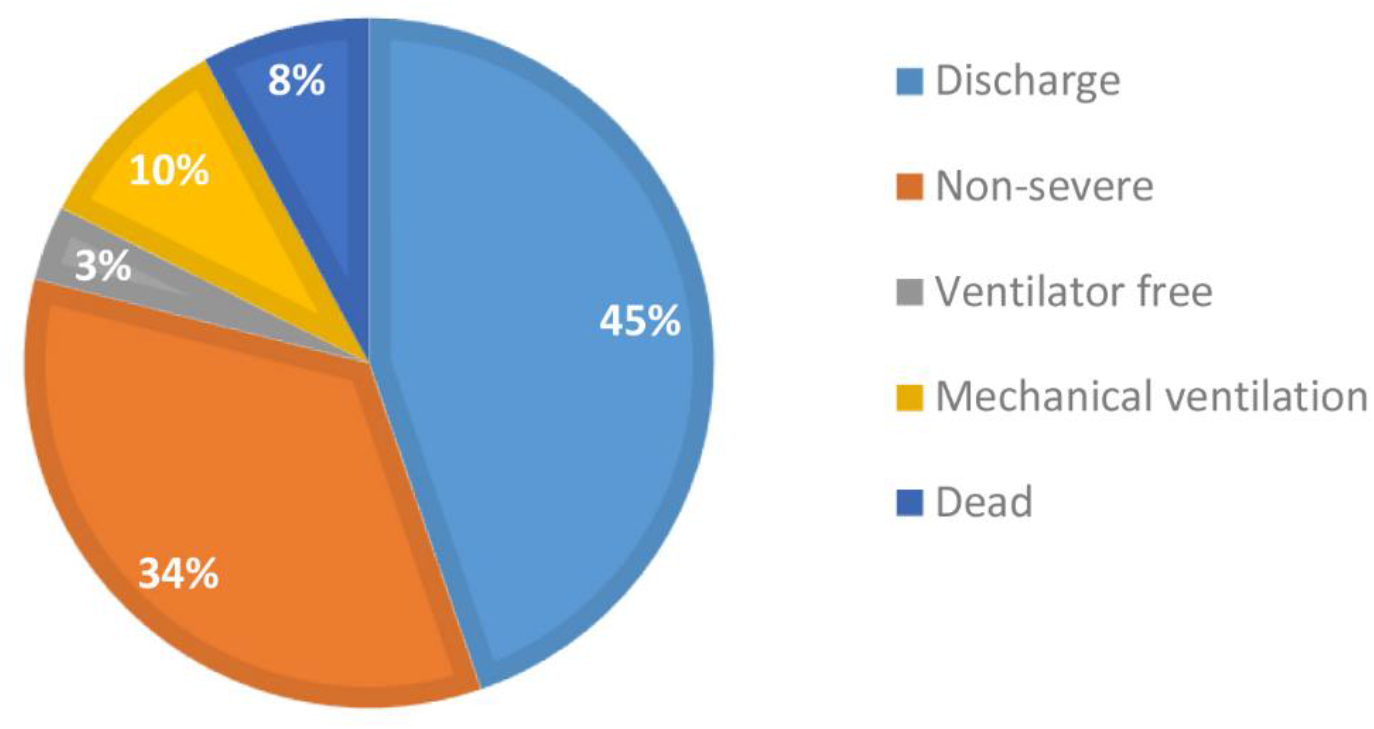
Distribution of short-term (28 days) outcomes of severe ill patients with COVID-19 (N=114).

### Demographics and Characteristics

General demographic and epidemiological characteristics of all enrolled patients were summarized in Table 1. The median age was 63.96 ± 13.41 years, and 58 (50.9%) were older than 65 years. Among the patients, 71 (62.3%) were male, 89 (78.1%) have chronic medical illness and the most coexisting disorders were hypertension 62 (54.4%), diabetes 39 (34.2%) and cardiovascular diseases 31 (27.2%). No significant differences were showed in such characteristics between outcome groups (*P* ≥ 0.05 for all). Table 2 displayed the clinical characteristics of the patients. The duration period from onset of symptoms to diagnosis with COVID-19 and severe ill was 4.0 (2.0-7.0) days and 10.0 (6.0-14.3) days, respectively. For 114 patients, the most common symptoms at initial were fever 78 (68.4%), cough 49 (43.0%), chest tightness 34 (29.8%), fatigue 30 (26.3%). Other symptoms including shortness of breath 18 (15.8%), anorexia 12 (10.5%), chill 12 (10.5%), mylgia 10 (8.8%), sputum 9 (7.9%), headache 8 (7.0%), diarrhea 8 (7.0%), chest pain 3 (2.6%), palpitation 3 (2.6%), stomachache 3 (2.6%), nausea 3 (2.6%), sore throat 2 (1.8%) were relatively rare. As for Oxygen saturation, patients in poor outcome group had significantly lower levels than that in good outcome group [median (IQR): 91 (90-93) % in good outcome group, 81 (74-88) % in bad outcome group, *P* < 0.001].

**Table 1.** Demographic and epidemiological characteristics of severe patients with COVID-19

|  | **Total (N=114)** | **Good outcome (N=94)** | **Poor outcome (N=20)** | **P** |
| --- | --- | --- | --- | --- |
| Age (years) | 63.96 ± 13.41 | 62.85 ± 13.65 | 69.15 ± 11.08 | 0.056 |
| < 65 | 52 (45.6) | 46 (48.9) | 6 (30.0) | 0.123 |
| ≥ 65 | 62 (54.4) | 48 (51.1) | 14 (70.0) |  |
| Sex, male | 71 (62.3) | 58 (61.7) | 13 (65.0) | 0.782 |
| Hospital infection | 7 (6.1) | 6 (6.4) | 1 (5.0) | > 0.999 |
| Coexisting disorders | 89 (78.1) | 73 (77.7) | 16 (80.0) | > 0.999 |
| Diabetes | 39 (34.2) | 34 (36.2) | 5 (25.0) | 0.339 |
| Hypertension | 62 (54.4) | 50 (53.2) | 12 (60.0) | 0.579 |
| Hyperlipidemia | 17 (14.9) | 15 (16.0) | 2 (10.0) | 0.739 |
| Cardiovascular diseases | 31 (27.2) | 24 (25.5) | 7 (35.0) | 0.388 |
| Cerebrovascular diseases | 6 (5.3) | 3 (3.2) | 3 (15.0) | 0.110 |
| Cancer | 10 (8.8) | 9 (9.6) | 1 (5.0) | 0.825 |
| Chronic renal diseases | 6 (5.3) | 4 (4.3) | 2 (10.0) | 0.622 |
| Chronic liver diseases | 4 (3.5) | 3 (3.2) | 1 (5.0) | 0.543 |
| Chronic Obstructive<br>Pulmonary Disease | 11 (9.6) | 9 (9.6) | 2 (10.0) | > 0.999 |
| Neuropsychiatric disorders | 3 (2.6) | 2 (2.1) | 1 (5.0) | 0.443 |
| History of surgery | 33 (28.9) | 26 (27.7) | 7 (35.0) | 0.511 |

Values are n (%) for categorical data, means ± SDs for normally distributed data, or medians (IQRs) for non-normally distributed data.

**Table 2.** Clinical characteristics of severe patients with COVID-19

|  | **Total**<br>**(N=114)** | **Good outcome**<br>**(N=94)** | **Poor outcome**<br>**(N=20)** | ***P*** |
| --- | --- | --- | --- | --- |
| Onset of symptom to, d |  |  |  |  |
| Dignosis | 4.0 (2.0-7.0) | 4.0 (2.0-7.0) | 4.5 (2.3-10.8) | 0.517 |
| Servious illness | 10.0 (6.0-14.3) | 10.0 (6.0-15.0) | 8.0 (5.0-14.0) | 0.540 |
| Signs and symptoms at<br>initial |  |  |  |  |
| Fever | 78 (68.4) | 63 (67.0) | 15 (75.0) | 0.486 |
| Chest tightness | 34 (29.8) | 27 (28.7) | 7 (35.0) | 0.577 |
| Shortness of breath | 18 (15.8) | 13 (13.8) | 5 (25.0) | 0.365 |
| Cough | 49 (43.0) | 39 (41.5) | 10 (50.0) | 0.485 |
| Sputum | 9 (7.9) | 8 (8.5) | 1 (5.0) | 0.943 |
| Fatigue | 30 (26.3) | 28 (29.8) | 2 (10.0) | 0.068 |
| Headache | 8 (7.0) | 7 (7.4) | 1 (5.0) | > 0.999 |
| Mylgia | 10 (8.8) | 8 (8.5) | 2 (10.0) | > 0.999 |
| Chest pain | 3 (2.6) | 3 (3.2) | 0 (0.0) | 1.000 |
| Anorexia | 12 (10.5) | 12 (12.8) | 0 (0.0) | 0.198 |
| Chill | 12 (10.5) | 8 (8.5) | 4 (20.0) | 0.263 |
| Stomachache | 3 (2.6) | 3 (3.2) | 0 (0.0) | > 0.999 |
| Diarrhea | 8 (7.0) | 7 (7.4) | 1 (5.0) | > 0.999 |
| Nausea | 3 (2.6) | 2 (2.1) | 1 (5.0) | 0.443 |
| Temperature at disease<br>onset (°C) | 38.1 (36.7-38.8) | 38.1 (36.7-38.7) | 38.2 (37.0-39.0) | 0.561 |
| < 37.4 | 36 (31.6) | 31 (33.0) | 5 (25.0) | 0.622 |
| 37.4-39.0 | 63 (55.3) | 50 (53.2) | 13 (65.0) |  |
| > 39.0 | 15 (13.2) | 13 (13.8) | 2 (10.0) |  |
| Signs and symptoms at<br>hospital admission |  |  |  |  |
| Fever | 32 (28.1) | 25 (26.6) | 7 (35.0) | 0.448 |
| Chest tightness | 50 (43.9) | 46 (48.9) | 4 (20.0) | 0.018 |
| Shortness of breath | 68 (59.6) | 49 (52.1) | 19 (95.0) | < 0.001 |
| Cough | 9 (7.9) | 7 (7.4) | 2 (10.0) | > 0.999 |
| Fatigue | 43 (37.7) | 37 (39.4) | 6 (30.0) | 0.433 |
| Headache | 6 (5.3) | 6 (6.4) | 0 (0.0) | 0.542 |
| Mylgia | 5 (4.4) | 5 (5.3) | 0 (0.0) | 0.585 |
| Chest pain | 3 (2.6) | 3 (3.2) | 0 (0.0) | > 0.999 |
| Anorexia | 18 (15.8) | 16 (17.0) | 2 (10.0) | 0.657 |
| Diarrhea | 10 (8.8) | 7 (7.4) | 3 (15.0) | 0.516 |
| Nausea | 2 (1.8) | 2 (2.1) | 0 (0.0) | > 0.999 |
| General signs at admission |  |  |  |  |
| Temperature (°C) | 36.7 (36.4-37.4) | 36.7 (36.4-37.3) | 38.2 (37.0-39.0) | 0.279 |
| < 37.4 | 85 (74.6) | 72 (76.6) | 13 (65.0) | 0.535 |
| 37.4-39.0 | 26 (22.8) | 20 (21.3) | 6 (30.0) |  |
| > 39.0 | 3 (2.6) | 2 (2.1) | 1 (5.0) |  |
| Heart rate (/min) | 86 (78-102) | 87 (78-102) | 88 (78-102) | 0.456 |
| < 100 | 83 (72.8) | 69 (73.4) | 14 (70.0) | 0.756 |
| > 100 | 31 (27.2) | 25 (26.6) | 6 (30.0) |  |
| Respiratory rate (/min) | 20 (20-23) | 20 (20-22) | 21 (20-30) | 0.122 |
| < 20 | 22 (19.3) | 19 (20.2) | 3 (15.0) | 0.760 |
| ≥ 20 | 92 (80.7) | 75 (79.8) | 17 (85.0) |  |
| Oxygen saturation (%) | 90 (88-92) | 91 (90-93) | 81 (74-88) | < 0.001 |
| < 90 | 35 (30.7) | 19 (20.2) | 16 (80.0) | < 0.001 |
| ≥ 90 | 79 (69.3) | 75 (79.8) | 4 (20.0) |  |

Values are n (%) for categorical data, means ± SDs for normally distributed data, or medians (IQRs) for non-normally distributed data.

### Laboratory findings and CT scans

Regarding to 114 severe patients, many laboratory indicators differed significantly between outcome groups (Table 3). Compare with good outcome group, the absolute counts of neutrophils 6.25 (4.69-9.20) *vs*. 3.48 (2.54-5.23), c-reactive protein 102.15 (78.07-122.90) *vs*. 48.95 (15.08-83.98), D dimer 2.10 (1.22-3.07) *vs*. 0.96 (0.41-1.78), total bilirubin 19.20 (9.25-33.05) *vs*. 11.05 (8.53-14.05), blood urea nitrogen 9.02 (5.26-11.30) *vs*. 4.11 (3.11- 5.04), creatine kinase 151.50 (50.50-218.50) *vs*. 62.00 (46.75-110.50), lactate dehydrogenase 638.00 (436.00-923.00) *vs*. 259.50 (213.75-382.50), hypersensitive cardiac troponin I 60.70 (18.48-298.98) *vs*. 4.10 (1.70-10.83), ferritin 679.00 (573.90-993.15) *vs*. 321.80 (231.00- 532.88), IL6 76.10 (19.05-192.88) *vs*. 21.23 (7.23-47.61), IL10 6.59 (4.58-11.78) *vs*. 4.64 (3.65-6.18) were significantly higher in poor outcome. Besides, total protein 60.40 (56.78- 64.05) *vs*. 63.80 (59.33-68.50), PaO2 68.15 (49.00-77.75) *vs*. 81.00 (74.75-89.00) were significantly lower in poor outcome group. For chest X- ray/CT, 107 (93.9) patients had Ground-glass opacity. These data indicated that the uncontrolled inflammation responses, infection, liver and kidney dysfunction and hypoxia may contribute to a poor outcome of COVID-19.

**Table 3.** Laboratory and radiographic findings at baseline of severe patients with COVID-19

|  | Total (N=114) | Good outcome (N=94) | Poor outcome (N=20) | P |
| --- | --- | --- | --- | --- |
| **Hematologic** |  |  |  |  |
| Leucocytes ($\times 10^9$ /L) | 6.28 ± 3.36 | 5.74 ± 2.49 | 8.78 ± 5.36 | 0.022 |
| Neutrophils ($\times 10^9$ /L) | 3.88 (2.65-5.81) | 3.48 (2.54-5.23) | 6.25 (4.69-9.20) | < 0.001 |
| Lymphocytes ($\times 10^9$ /L) | 0.87 (0.65-1.31) | 0.92 (0.70-1.43) | 0.67 (0.43-0.89) | 0.001 |
| Monocytes ($\times 10^9$ /L) | 0.44 (0.30-0.59) | 0.48 (0.33-0.61) | 0.34 (0.22-0.40) | 0.009 |
| Platelets ($\times 10^9$ /L) | 192.00 (140.74-269.75) | 205.00 (142.75-272.50) | 165.00 (138.25-218.00) | 0.160 |
| Haemoglobin (g/L) | 122.04 ± 17.78 | 122.65 ± 16.75 | 119.15 ± 22.29 | 0.427 |
| CD4+ T cells (%) | 43.54 (36.45-53.41) | 44.98 (36.53-53.20) | 39.95 (31.46-57.32) | 0.687 |
| CD8+ T cells (%) | 21.53 (16.48-28.55) | 22.11 (17.61-29.95) | 17.90 (14.00-23.45) | 0.025 |
| **Coagulation function** |  |  |  |  |
| Prothrombin time (s) | 13.5 (12.8-14.3) | 13.40 (12.70-14.23) | 14.25 (12.93-15.28) | 0.072 |
| Activated partial thromboplastin time (s) | 38.00 (35.18-41.53) | 38.00 (35.10-41.43) | 38.45 (35.45-43.98) | 0.398 |
| Fibrinogen (g/L) | 4.98 ± 1.48 | 5.05 ± 1.43 | 4.66 ± 1.67 | 0.282 |
| Thrombin time (s) | 17.35 (16.58-18.30) | 17.35 (16.77-18.15) | 17.20 (15.35-19.18) | 0.467 |
| D-dimer (mg/L) | 1.06 (0.51-2.10) | 0.96 (0.41-1.78) | 2.10 (1.22-3.07) | 0.005 |
| **Biochemical** |  |  |  |  |
| Alanine aminotransferase (U/L) | 45.50 (26.00-74.25) | 44.50 (25.75-72.50) | 49.00 (29.25-80.25) | 0.427 |
| Aspartate aminotransferase (U/L) | 39.50 (26.75-64.50) | 38.50 (26.00-57.25) | 44.00 (29.50-114.75) | 0.197 |
| Total bilirubin (μmol/L) | 11.35 (8.93-16.15) | 11.05 (8.53-14.05) | 19.20 (9.25-33.05) | 0.024 |
| Total protein (g/L) | 63.10 (58.48-67.70) | 63.80 (59.33-68.50) | 60.40 (56.78-64.05) | 0.015 |
| Albumin (g/L) | 34.65 (30.350-38.60) | 35.80 (31.85-39.10) | 30.05 (27.10-32.65) | < 0.001 |
| Globulin (g/L) | 29.15 (25.70-31.65) | 28.95 (25.68-30.78) | 30.35 (29.63-35.50) | 0.057 |
| Prealbumin (mg/L) | 120 (90-153) | 120 (90-151) | 93 (82-181) | 0.145 |
| Blood urea nitrogen (mmol/L) | 4.39 (3.27-6.13) | 4.11 (3.11-5.04) | 9.02 (5.26-11.30) | < 0.001 |
| Creatinine (μmol/L) | 74.35 (60.00-87.85) | 74.35 (60.28-86.38) | 71.80 (53.70-95.60) | 0.636 |
| Creatine kinase (U/L) | 66.50 (46.75-133.50) | 62.00 (46.75-110.50) | 151.50 (50.50-218.50) | 0.046 |
| Creatine kinase-MB (U/L) | 17.00 (14.00-23.00) | 17.00 (14.00-22.00) | 19.50 (13.00-31.75) | 0.234 |
| Lactate dehydrogenase (U/L) | 286.00 (223.75-452.25) | 259.50 (213.75-382.50) | 638.00 (436.00-923.00) | < 0.001 |
| Hypersensitive cardiac troponin I (ng/L) | 5.70 (2.10-19.05) | 4.10 (1.70-10.83) | 60.70 (18.48-298.98) | < 0.001 |
| Glucose (mmol/L) | 6.20 (5.20-8.31) | 6.13 (5.16-7.63) | 8.26 (5.81-13.42) | 0.013 |
| Serum potassium (mmol/L) | 4.03 (3.70-4.50) | 4.07 (3.72-4.51) | 3.97 (3.59-4.36) | 0.344 |
| Serum sodium (mmol/L) | 138.85 (136.30-142.00) | 138.20 (136.00-141.68) | 142.40 (138.85-146.93) | 0.001 |
| Serum calcium (mmol/L) | 2.15 ± 0.18 | 2.17 ± 0.16 | 2.04 ± 0.20 | 0.003 |
| Serum phosphorus (mmol/L) | 1.00 (0.88-1.11) | 1.00 (0.89-1.11) | 0.95 (0.73-1.12) | 0.381 |
| Serum chlorine (mmol/L) | 100.48 ± 5.10 | 100.03 ± 4.45 | 102.61 ± 7.20 | 0.137 |
| Lactate concentration (mmol/L) | 1.60 (1.38-1.80) | 1.50 (1.30-1.70) | 2.10 (1.70-2.40) | < 0.001 |
| Positive Urinary protein, n | 47 (41.2) | 33 (35.1) | 14 (70.0) | 0.004 |
| Positive Urinary glucose, n | 8 (7.0) | 7 (7.4) | 1 (5.0) | > 0.999 |
| Positive urinary occult blood, n | 31 (27.2) | 21 (22.3) | 10 (50.0) | 0.012 |
| **Blood gas characteristics** |  |  |  |  |
| pH | 7.39 (7.36-7.42) | 7.38 (7.36-7.42) | 7.41 (7.31-7.46) | 0.636 |
| Arterial partial pressure of oxygen (mm Hg) | 79.00 (70.00-88.00) | 81.00 (74.75-89.00) | 68.15 (49.00-77.75) | < 0.001 |
| Arterial partial pressure of carbon dioxide (mm Hg) | 43.00 (38.00-46.00) | 42.80 (38.00-45.00) | 45.50 (35.60-57.00) | 0.198 |
| **Infection-related biomarkers** |  |  |  |  |
| Interleukin 2 (pg/mL) | 2.70 (2.43-3.04) | 2.70 (2.47-3.02) | 2.69 (2.41-3.60) | 0.991 |
| Interleukin 4 (pg/mL) | 2.16 (1.85-2.60) | 2.16 (1.89-2.52) | 2.21 (1.68-3.50) | 0.729 |
| Interleukin 6 (pg/mL) | 23.28 (8.31-54.23) | 21.23 (7.23-47.61) | 76.10 (19.05-192.88) | 0.002 |
| Interleukin 10 (pg/mL) | 4.91 (3.92-6.74) | 4.64 (3.65-6.18) | 6.59 (4.58-11.78) | 0.001 |
| Tumour necrosis factor-α (pg/mL) | 2.63 (2.11-4.80) | 2.63 (2.11-4.80) | 2.67 (2.07-4.66) | 0.838 |
| Interferon-γ (pg/mL) | 2.50 (1.96-3.20) | 2.46 (1.96-3.20) | 2.58 (1.89-3.49) | 0.571 |
| C-reactive protein (mg/L) | 67.95 (20.50-103.25) | 48.95 (15.08-83.98) | 102.15 (78.07-122.90) | < 0.001 |
| Ferritin (ng/mL) | 390.60 (261.70-721.00) | 321.80 (231.00-532.88) | 679.00 (573.90-993.15) | 0.001 |
| IgM, n | 60 (68.2) | 52 (69.3) | 8 (61.5) | 0.748 |
| IgG, n | 88 (100.0) | 75 (100.0) | 13 (100.0) | NA |
| **Chest X- ray/CT findings** |  |  |  |  |
| Ground-glass opacity | 107 (93.9) | 87 (92.6) | 20 (100.0) | 0.351 |
| Unilateral pneumonia | 11 (9.6) | 11 (11.7) | 0 (0.0) | 0.208 |
| Bilateral pneumonia | 102 (89.5) | 82 (87.2) | 20 (100.0) | 0.122 |
| Interstitial abnormalities | 21 (18.4) | 15 (16.0) | 6 (30.0) | 0.200 |

Data are n (%), means ± SDs or medians (IQRs) when appropriate.

**Table 4.** Complications and treatments of severe patients with COVID-19

|  | **Total**<br>**(N=114)** | **Good outcome**<br>**(N=94)** | **Poor outcome**<br>**(N=20)** | ***P*** |
| --- | --- | --- | --- | --- |
| **Complications** |  |  |  |  |
| Shock | 8 (7.0) | 0 (0.0) | 8 (40.0) | < 0.001 |
| Acute respiratory distress syndrome | 41 (36.0) | 21 (22.3) | 20 (100.0) | < 0.001 |
| Acute renal injury | 35 (30.7) | 21 (22.3) | 14 (70.0) | < 0.001 |
| Acute myocardial injury | 28 (24.6) | 13 (13.8) | 15 (75.0) | < 0.001 |
| Acute liver function injury | 69 (60.5) | 57 (60.6) | 12 (60.0) | 0.958 |
| Arrhythmia | 31 (27.2) | 16 (17.0) | 15 (75.0) | < 0.001 |
| Rhabdomyolysis | 11 (9.6) | 2 (2.1) | 9 (45.0) | < 0.001 |
| Disseminated intravascular coagulation | 15 (13.2) | 2 (2.1) | 13 (65.0) | < 0.001 |
| **Treatment** |  |  |  |  |
| Antibiotic treatment | 114 (100.0) | 94 (100.0) | 20 (100.0) | NA |
| Anticoronavirus treatment | 113 (99.1) | 93 (98.9) | 20 (100.0) | > 0.999 |
| Glucocorticoids | 47 (41.2) | 28 (29.8) | 19 (95.0) | < 0.001 |
| Oxygen therapy | 114 (100.0) | 94 (100.0) | 20 (100.0) | NA |
| Immunoglobulin | 64 (56.1) | 45 (47.9) | 19 (95.0) | < 0.001 |
| Parenteral nutrition | 49 (43.0) | 29 (30.9) | 20 (100.0) | < 0.001 |
| Admission to intensive care unit | 29 (25.4) | 9 (9.6) | 20 (100.0) | < 0.001 |
| Noninvasive ventilation | 25 (21.9) | 13 (13.8) | 12 (60.0) | < 0.001 |
| Invasive mechanical ventilation | 22 (19.3) | 4 (4.3) | 18 (90.0) | < 0.001 |
| Extracorporeal membrane oxygenation | 6 (5.3) | 0 (0.0) | 6 (30.0) | < 0.001 |
| Vasoconstrictive agents | 20 (17.5) | 1 (1.1) | 19 (95.0) | < 0.001 |
| Renal replacement therapy | 2 (1.8) | 0 (0.0) | 2 (10.0) | 0.029 |
| Traditional Chinese medicine | 86 (75.4) | 77 (81.9) | 9 (45.0) | 0.001 |
| Trastuzumab | 13 (11.4) | 12 (12.8) | 1 (5.0) | 0.459 |
| Infusions of blood plasma | 4 (3.5) | 2 (2.1) | 2 (10.0) | 0.141 |
| **Onset of severe illness to, d\*** |  |  |  |  |
| 1<sup>st</sup> for RT-PCR (-) | 14.0 (11.0-18.0) | 14.0 (10.0-18.0) | 19.0 (15.5-24.0) | <0.001 |
| 2<sup>nd</sup> for RT-PCR (-) | 17.5 (14.0-22.0) | 17.0 (14.0-21.5) | 24.0 (22.0-27.5) | <0.001 |

Data are n (%) or medians (IQRs) when appropriate.
\*Data available for 98 patients.

### Complications and treatments

The severe patients with COVID-19 had complications of acute liver injury, ARDS, acute kidney injury, arrhythmia, acute myocardial injury, DIC, rhabdomyolysis, septic shock and among them, nobody had septic shock in good outcome group. Otherwise, in poor outcome group, all patients had ARDS. Acute myocardial injury, acute kidney injury, arrhythmia, rhabdomyolysis, DIC were significantly higher than their counterparts of 13.8 %, 22.3 %, 17.0%, 2.1% and 2.1% in patients with COVID-19 in good outcome group, respectively. All 114 patients with COVID-19 were treated with antibiotics and high flow nasal cannula, 25 (21.9%) with non-invasive mechanical ventilation and 22 (19.3%) with invasive mechanical ventilation treatment. Six (5.3%) patients were treated with extracorporeal membrane oxygenation (ECMO) and all of them end up with poor outcome. Almost all [113 (99.1%)] patients received antiviral treatment including the arbidol hydrochloride capsules (0.2 g three times daily), lopinavir and ribavirin (500 mg two times daily) via the oral route. Furthermore, as many as 41.2% patients received glucocorticoid therapy. Sixty-four (56.1%) patients received immunoglobulin treatment, 49 (43.0%) patients were treated with parenteral nutrition, the percentage was higher in poor outcome group than in good outcome group [20 (100.0%) *vs*. 29 (30.9%)]. Two patients (1.8 %) were treated with renal replacement therapy, and 20 (17.5%) with vasoconstrictive agents and it was higher than in good outcome group [19 (95.0%) *vs*. 1 (1.1%)]. All 20 poor outcome patients were transferred to the ICU, which was significantly higher than that of 29 (25.4%) in the good outcome group.

### Prediction of risk factors for severe COVID-19 in poor outcome group

Table 5-6 displayed the results of univariate and multivariate Cox analyses of potential factors for short-term outcomes in severe patients with COVID-19. Our results indicated that, for severe patients, higher levels of oxygen saturation (HR, 0.123; 95% CI, 0.041-0.369), albumin (HR, 0.060; 95% CI, 0.008-0.460) and arterial partial pressure of oxygen (HR, 0.321; 95% CI, 0.106-0.973) were associated with decreased risk of developing poor outcome within 28 days. In the other hand, higher levels of leucocytes (HR, 5.575; 95% CI, 2.080- 14.943), neutrophils (HR, 2.566; 95% CI, 1.022-6.443), total bilirubin (HR, 6.171; 95% CI, 2.458-15.496), globulin (HR, 2.526; 95% CI, 1.027-6.211), blood urea nitrogen (HR, 5.640; 95% CI, 2.193-14.509), creatine kinase-MB (HR, 3.032; 95% CI, 1.203-7.644), lactate dehydrogenase (HR, 4.607; 95% CI, 1.057-20.090), hypersensitive cardiac troponin I (HR, 5.023; 95% CI, 1.921-13.136), lactate concentration (HR,15.721; 95% CI, 2.099-117.777), Interleukin-10 (HR, 3.551; 95% CI, 1.280-9.857) and C-reactive protein (HR, 5.275; 95% CI, 1.517-18.344) were associated with increased risk of poor outcome development. For all the factors analyzed above, increased concentration of lactate (≥ 1.6mmol/L) and total bilirubin (≥ 19.0 μmol/L) might be the most important predictors of poor outcome in the early stage. Furthermore, we found that there was a nonlinear dose-response relationship between ten indices and poor outcome risk, which is depicted in Figure 2.

**Table 5.** Univariate and multivariate analyses of potential factors (demographic and epidemiologic) predicting poor outcome

| Factors | Level | Crude HR (95% CI) | *P* | Adjusted HR (95% CI)* | *P* |
| --- | --- | --- | --- | --- | --- |
| Age (years) | ≥ 65 v.s. < 65 | 2.192 (0.842-5.708) | 0.108 | 2.184 (0.839-5.687) | 0.110 |
| Sex | Female v.s. Male | 0.772 (0.308-1.937) | 0.581 | 0.732 (0.292-1.838) | 0.507 |
| Coexisting disorders | Yes v.s. No | 1.154 (0.386-3.453) | 0.797 | 0.692 (0.207-2.313) | 0.550 |
| Diabetes | Yes v.s. No | 0.706 (0.257-1.945) | 0.380 | 0.622 (0.224-1.728) | 0.363 |
| Hypertension | Yes v.s. No | 1.122 (0.458-2.747) | 0.801 | 0.960 (0.386-2.384) | 0.929 |
| Hyperlipidemia | Yes v.s. No | 0.677 (0.157-2.919) | 0.601 | 0.729 (0.168-3.167) | 0.673 |
| Cardiovascular diseases | Yes v.s. No | 1.601 (0.638-4.015) | 0.316 | 1.062 (0.380-2.970) | 0.908 |
| Cerebrovascular diseases | Yes v.s. No | 3.327 (0.975-11.356) | 0.055 | 2.326 (0.612-8.848) | 0.216 |
| Cancer | Yes v.s. No | 0.536 (0.072-4.004) | 0.543 | 0.410 (0.054-3.103) | 0.388 |
| Chronic renal diseases | Yes v.s. No | 3.678 (0.835-16.202) | 0.085 | 3.437 (0.764-15.465) | 0.108 |
| Chronic liver diseases | Yes v.s. No | 1.433 (0.192-10.707) | 0.726 | 0.997 (0.128-7.760) | 0.997 |
| Chronic Obstructive Pulmonary Disease | Yes v.s. No | 0.991 (0.230-4.272) | 0.990 | 0.642 (0.139-2.955) | 0.569 |
| Neuropsychiatric disorders | Yes v.s. No | 1.186 (0.158-8.894) | 0.868 | 0.734 (0.094-5.741) | 0.768 |
| History of surgery | Yes v.s. No | 1.168 (0.466-2.930) | 0.740 | 1.041 (0.413-2.623) | 0.932 |
| Temperature at disease onset (°C) | 37.4-39.0 v.s. < 37.4 | 1.514 (0.540-4.248) | 0.430 | 1.844 (0.638-5.326) | 0.258 |
|  | > 39.0 v.s. < 37.4 | 1.036 (0.201-5.342) | 0.966 | 2.714 (0.348-21.190) | 0.622 |
| Temperature at admission (°C) | 37.4-39.0 v.s. < 37.4 | 1.476 (0.561-3.885) | 0.430 | 1.721 (0.646-4.580) | 0.277 |
|  | > 39.0 v.s. < 37.4 | 2.630 (0.343-20.167) | 0.352 | 2.714 (0.348-21.190) | 0.341 |
| Respiratory rate (/min) | ≥ 20 v.s. < 20 | 1.357 (0.398-4.629) | 0.626 | 1.316 (0.385-4.502) | 0.662 |
| Oxygen saturation (%) | ≥ 90 v.s. < 90 | 0.131 (0.044-0.394) | < 0.001 | 0.123 (0.041-0.369) | < 0.001 |

\*Adjustments were made for age and sex.

**Table 6.** Univariate and multivariate analyses of potential factors (laboratory indexes) predicting poor outcome

| Factors | Normal range | Level<sup>†</sup> | Crude HR (95% CI) | P | Adjusted HR (95% CI)* | P |
| --- | --- | --- | --- | --- | --- | --- |
| **Hematologic** |  |  |  |  |  |  |
| Leucocytes (×10<sup>9</sup> /L) | 3.5-9.5 | ≥ 9.5 v.s. < 9.5 | 4.634 (1.840-11.669) | 0.001 | 5.575 (2.080-14.943) | 0.001 |
| Neutrophils (×10<sup>9</sup> /L) | 1.8-6.3 | ≥ 6.3 v.s. < 6.3 | 2.663 (1.102-6.433) | 0.030 | 2.566 (1.022-6.443) | 0.045 |
| Lymphocytes (×10<sup>9</sup> /L) | 1.1-3.2 | ≥ 1.1 v.s. < 1.1 | 0.293 (0.068-1.266) | 0.100 | 0.337 (0.077-1.475) | 0.149 |
| Monocytes (×10<sup>9</sup> /L) | 0.1-0.6 | ≥ 0.6 v.s. < 0.6 | 0.179 (0.024-1.335) | 0.179 | 0.182 (0.024-1.366) | 0.098 |
| Platelets (×10<sup>9</sup> /L) | 125.0-350.0 | ≥ 125.0 v.s. < 125.0 | 0.733 (0.245-2.192) | 0.578 | 0.837 (0.275-2.553) | 0.755 |
| Haemoglobin (g/L) | 130.0-175.0 | ≥ 130.0 v.s. < 130.0 | 0.865 (0.354-2.116) | 0.751 | 0.652 (0.244-1.732) | 0.394 |
| CD4+ T cells (%) | 25.34-51.37 | ≥ 51.37 v.s. < 51.37 | 1.144 (0.439-2.980) | 0.783 | 1.235 (0.468-3.259) | 0.670 |
| CD8+ T cells (%) | 14.23-38.95 | ≥ 14.23 v.s. < 14.23 | 0.687 (0.249-1.890) | 0.467 | 0.867 (0.303-2.482) | 0.790 |
| **Coagulation function** |  |  |  |  |  |  |
| Prothrombin time (s) | 11.0-16.0 | ≥ 13.5 v.s. < 13.5 | 1.574 (0.644-3.852) | 0.320 | 1.112 (0.417-2.966) | 0.832 |
| Activated partial thromboplastin time (s) | 28.0-43.5 | ≥ 43.5 v.s. < 43.5 | 1.363 (0.493-3.766) | 0.551 | 1.204 (0.433-3.344) | 0.722 |
| Fibrinogen (g/L) | 2.0-4.0 | ≥ 4.0 v.s. < 4.0 | 0.533 (0.212-1.338) | 0.180 | 0.520 (0.205-1.317) | 0.168 |
| Thrombin time (s) | 14.0-21.0 | ≥ 17.4 v.s. < 17.4 | 1.114 (0.463-2.678) | 0.809 | 1.197 (0.489-2.930) | 0.694 |
| D-dimer (mg/L) | < 0.5 | ≥ 0.5 v.s. < 0.5 | 1.232 (0.411-3.697) | 0.710 | 0.940 (0.294-3.001) | 0.917 |
| **Biochemical** |  |  |  |  |  |  |
| Alanine aminotransferase (U/L) | 5-40 | ≥ 40 v.s. < 40 | 0.961 (0.393-2.351) | 0.931 | 1.203 (0.429-3.373) | 0.726 |
| Aspartate aminotransferase (U/L) | 8-40 | ≥ 40 v.s. < 40 | 1.611 (0.658-3.942) | 0.296 | 1.900 (0.755-4.783) | 0.173 |
| Total bilirubin (μmol/L) | 5.1-19.0 | ≥ 19.0 v.s. < 19.0 | 5.849 (2.433-14.063) | < 0.001 | 6.171 (2.458-15.496) | < 0.001 |
| Total protein (g/L) | 60-80 | ≥ 60 v.s. < 60 | 0.687 (0.284-1.661) | 0.405 | 0.721 (0.298-1.748) | 0.470 |
| Albumin (g/L) | 35-55 | ≥ 35 v.s. < 35 | 0.054 (0.007-0.405) | 0.054 | 0.060 (0.008-0.460) | 0.007 |
| Globulin (g/L) | 20-30 | ≥ 30 v.s. < 30 | 2.723 (1.113-6.666) | 0.028 | 2.526 (1.027-6.211) | 0.043 |
| Prealbumin (mg/L) | 170-420 | ≥ 170 v.s. < 170 | 1.001 (0.364-2.755) | 0.999 | 1.282 (0.448-3.665) | 0.643 |
| Blood urea nitrogen (mmol/L) | 2.9-8.2 | ≥ 8.2 v.s. < 8.2 | 6.283 (2.565-15.391) | < 0.001 | 5.640 (2.193-14.509) | < 0.001 |
| Creatinine (μmol/L) | 44-133 | ≥ 74 v.s. < 74 | 0.973 (0.405-2.337) | 0.950 | 0.709 (0.254-1.978) | 0.511 |
| Creatine kinase (U/L) | 38-174 | ≥ 174 v.s. < 174 | 2.039 (0.783-5.307) | 0.144 | 1.982 (0.756-5.199) | 0.164 |
| Creatine kinase-MB (U/L) | 0-24 | ≥ 24 v.s. < 24 | 2.449 (1.000-5.997) | 0.050 | 3.032 (1.203-7.644) | 0.019 |
| Lactate dehydrogenase (U/L) | 109-245 | ≥ 245 v.s. < 245 | 3.963 (0.915-17.161) | 0.066 | 4.607 (1.057-20.090) | 0.042 |
| Hypersensitive cardiac troponin I (ng/L) | < 26.2 | ≥ 26.2 v.s. < 26.2 | 5.613 (2.233-14.112) | < 0.001 | 5.023 (1.921-13.136) | 0.001 |
| Glucose (mmol/L) | 3.9-6.1 | ≥ 6.1 v.s. < 6.1 | 1.678 (0.637-4.416) | 0.295 | 1.454 (0.543-3.893) | 0.457 |
| Serum potassium (mmol/L) | 3.5-5.2 | ≥ 4.0 v.s. < 4.0 | 1.112 (0.462-2.677) | 0.813 | 0.921 (0.368-2.304) | 0.860 |
| Serum sodium (mmol/L) | 136-145 | ≥ 136 v.s. < 136 | 1.881 (0.436-8.120) | 0.397 | 3.302 (0.702-15.535) | 0.131 |
| Serum calcium (mmol/L) | 2.03-2.54 | ≥ 2.03 v.s. < 2.03 | 0.385 (0.160-0.925) | 0.033 | 0.433 (0.173-1.083) | 0.073 |
| Serum phosphorus (mmol/L) | 0.96-1.62 | ≥ 0.96 v.s. < 0.96 | 0.831 (0.345-2.001) | 0.680 | 0.794 (0.330-1.914) | 0.608 |
| Serum chlorine (mmol/L) | 96-108 | ≥ 96 v.s. < 96 | 1.617 (0.375-6.970) | 0.519 | 2.195 (0.489-9.845) | 0.305 |
| Lactate concentration (mmol/L) | 0.5-1.6 | ≥ 1.6 v.s. < 1.6 | 15.457 (2.067-115.615) | 0.008 | 15.721 (2.099-117.777) | 0.007 |
| Positive Urinary protein | / | / | 3.239 (1.244-8.433) | 0.016 | 2.905 (1.099-7.678) | 0.032 |
| Positive Urinary glucose | / | / | 0.893 (0.120-6.676) | 0.912 | 0.961 (0.128-7.238) | 0.969 |
| Positive urinary occult blood | / | / | 2.474 (1.030-5.945) | 0.043 | 2.247 (0.932-5.421) | 0.071 |
| **Blood gas characteristics** |  |  |  |  |  |  |
| pH | 7.35-7.45 | ≥ 7.35 v.s. < 7.35 | 0.427 (0.170-1.074) | 0.071 | 0.468 (0.181-1.207) | 0.116 |
| Arterial partial pressure of oxygen (mm Hg) | 80-100 | ≥ 80 v.s. < 80 | 0.295 (0.098-0.883 ) | 0.029 | 0.321 (0.106-0.973) | 0.045 |
| Arterial partial pressure of carbon dioxide (mm Hg) | 35-45 | ≥ 45 v.s. < 45 | 2.159 (0.892-5.230) | 0.088 | 2.224 (0.895-5.525) | 0.085 |

| **Infection-related biomarkers** |  |  |  |  |  |  |
| --- | --- | --- | --- | --- | --- | --- |
| Interleukin-2 (pg/mL) | 0.1-4.1 | $\geq 4.1$ v.s. $< 4.1$ | 1.820 (0.533-6.212) | 0.339 | 2.343 (0.654-8.389) | 0.191 |
| Interleukin-4 (pg/mL) | 0.1-3.2 | $\geq 3.2$ v.s. $< 3.2$ | 1.663 (0.663-4.172) | 0.279 | 2.112 (0.797-5.597) | 0.133 |
| Interleukin-6 (pg/mL) | 0.1-2.9 | $\geq 23.3$ v.s. $< 23.3$ | 1.782 (0.711-4.468) | 0.218 | 1.485 (0.575-3.836) | 0.414 |
| Interleukin-10 (pg/mL) | 0.1-5.0 | $\geq 5.0$ v.s. $< 5.0$ | 2.629 (1.010-6.843) | 0.048 | 3.551 (1.280-9.857) | 0.015 |
| Tumour necrosis factor-α (pg/mL) | 0.1-23.0 | $\geq 2.6$ v.s. $< 2.6$ | 1.079 (0.448-2.600) | 0.866 | 1.032 (0.427-2.491) | 0.945 |
| Interferon-γ (pg/mL) | 0.1-18.0 | $\geq 2.5$ v.s. $< 2.5$ | 1.260 (0.522-3.043) | 0.607 | 1.421 (0.582-3.473) | 0.440 |
| C-reactive protein (mg/L) | $< 8.0$ | $\geq 65.0$ v.s. $< 65.0$ | 4.703 (1.374-16.093) | 0.014 | 5.275 (1.517-18.344) | 0.009 |
| Ferritin (ng/mL) | 4.6-204.0 | $\geq 204.0$ v.s. $< 204.0$ | 2.582 (0.210-11.930) | 0.657 | 2.647 (0.311-22.522) | 0.373 |

\*Adjustments were made for age and sex.
†Cut points of levels were determined according to normal range, actual distribution and clinical significance.

**Figure 2.**
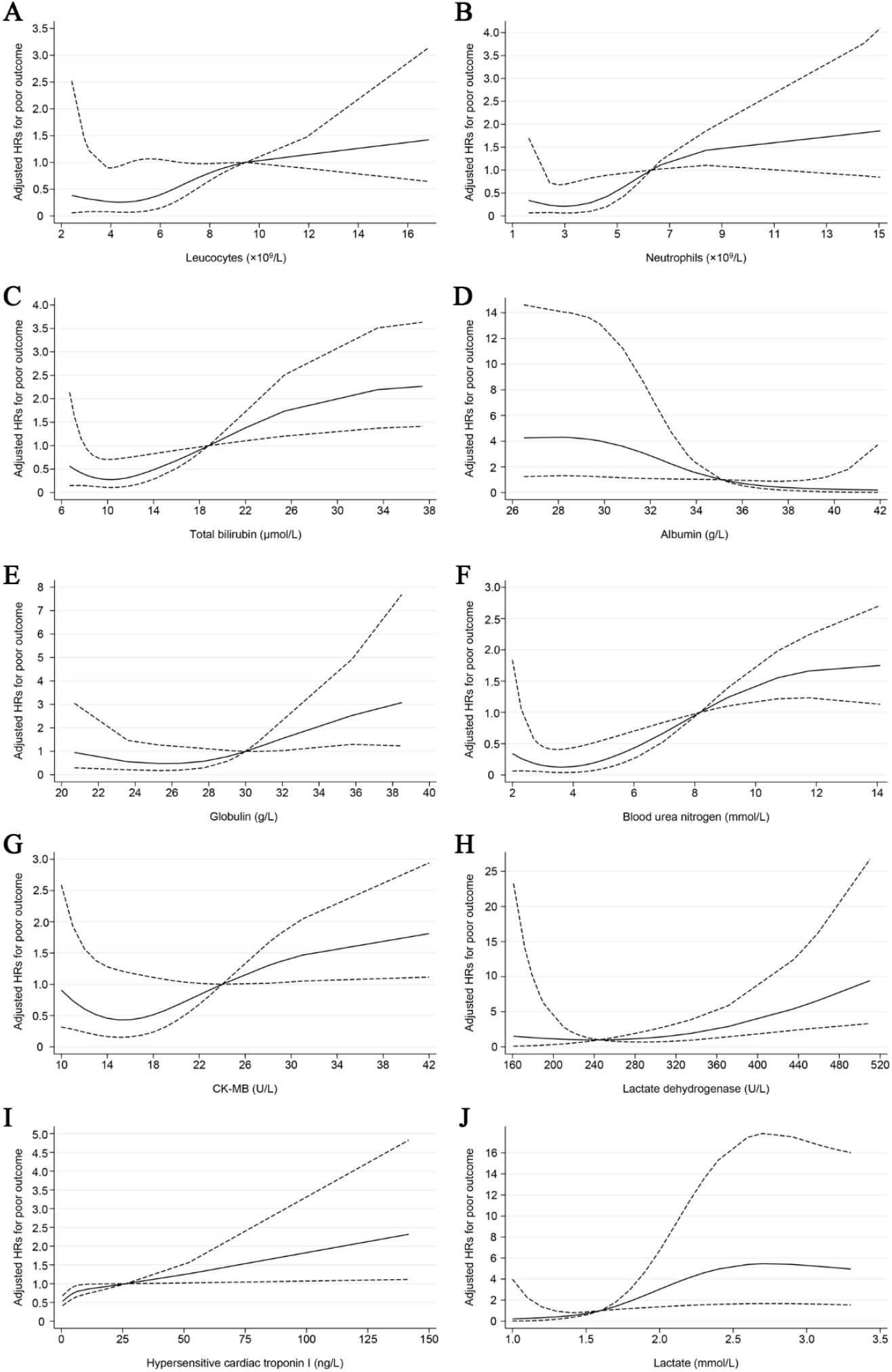
Nonlinear dose-response relationship between ten indices and poor outcome risk. Hazard ratios (HRs) were adjusted for age and gender. Dotted lines represent the 95% CIs for the fitted trend. (A) Leucocytes (×10^9^ /L), referent (HR =1): 9.5; (B) Neutrophils (×10^9^ /L), referent: 6.3; (C) Total bilirubin (μmol/L), referent: 19.0; (D) Albumin, referent: 35.0 (g/L); (E) Globulin (g/L), referent: 30.0; (F)Blood urea nitrogen (mmol/L), referent: 8.2; (G) CK- MB (U/L), referent: 24.0; (H) Lactate dehydrogenase (U/L), referent: 245; (I) Hypersensitive cardiac troponin I (ng/L), referent: 26.2; (J)Lactate (mmol/L), referent: 1.6.

## Discussion

SARS-CoV-2 has now spread globally and seriously threaten the human health. Demographics and Characteristics of COVID-19 has been gradually investigated and explored by many scientists. Studies had shown that 20% of COVID-19 patients developed to critically disease due to hypoxia or respiratory failure, among them, 5% needed to treat in intensive care unit and 15% required oxygen and essential care. This suggests that this is particularly important in understanding this part of the patient.[9] [10] Recently, Dong et al found that children, in particular, infants developed into severe outcomes. This indicated that patients of any age could develop to severe illness.[11] Feng et al found that severe and critical patients with the typical characteristics of multiple organ and immune function dysfunction. They also found that older people with age (≥ 75 years) was a risk factor for mortality.[12] With the increase number of asymptomatic infectious patients, take measures to detect and isolate early are especially important. In our study, we take a short-term method to prospectively study the reported the epidemiology and risk factors of 114 severe patients with COVID-19 from Union hospital, Hubei province. To our knowledge, this is the first report to describes the severe patients with COVID-19 during a short-term observation and predicted some risk factors for final outcome. In our study, the average age of severe patients swere 63.96 ± 13.41 years and 58 (50.9%) were older than 65 years, the patients are older than in other studies.[5, 13, 14] We also found that 78 (68.4%) of 114 patients were had fever at initial, this was in accordance with previous studies, which fever is the one of the most common symptom in patients who had COVID-19.[5, 14-16] But, 36 (31.6%) of 114 severe patients who were had not fever at the beginning of illness, therefore, other clinical manifestations should be concerned. In recently, Jin et al found that attention should also pay on people who had gastrointestinal symptoms.[6] Mao et al indicated that patients with Neurologic manifestations, clinicians should suspect COVID-19.[14]

According to results from laboratory tests, poor outcome group had lower lymphocytes than good outcome group [0.67 (0.43-0.89) *vs*. 0.92 (0.70-1.43)]. As is known to all, lymphocytes are the main fighting force against the virus, we suspected that SARS-CoV-2 viral damages on the lymphocyte and causes its reduction.[17] Chen et al found that severe lymphopenia were persistent and more in dead patients than recovered patients and they suggested that a lymphopenia may associated with poor outcome.[18] Tan et al demonstrated a oppose result that lymphopenia is an effective indicator for the severity of patients with COVID-19.[19] CD8+ T cells were significantly lower in poor outcome group. Chen et al indicated that the SARS-CoV-2 infection may affect CD4+ and CD8+ T lymphocytes cells particularly and argue that this is potential correlation with COVID-19 severity.[20] In addition, markedly higher concentrations of cardiac troponin I, creatine kinase, lactate dehydrogenase could be observed in poor outcome group than in their counterpart. Most notably, patients who had poor outcome may develop pulmonary and extra-pulmonary organ damage, including septic shock, acute respiratory distress syndrome, acute kidney injury, acute cardiac injury as well as disseminated intravascular coagulation. Fatality risk of COVID-19 patients with or without history of previous cardiovascular disease may the acute cardiac injury and heart failure.[18] Costanza Emanueli et al suggested that the COVID-19 crisis will have a longer term residual repercussions on the cardiovascular system.[21] This suggestion that the cardiac injury also requires special attention. In our study, we also found that lactate concentration was higher in poor outcome than their counterpart. As with all known, lactate is generally the end product of energy through anaerobic metabolism, and the elevation of lactate level is mainly caused by the increase of blood oxygen deficiency and anaerobic metabolism, this result is consistent with the lower oxygen saturation in poor group, this indicated that lactate level was an important predictors of poor outcome in the early stage. In addition, total bilirubin was also an important predictor of poor outcome in the early stage. Qi recommend that dynamic monitor the liver function of patients is necessary.[22] Cai conclude that patients with abnormal liver function may had higher risks to progress to severe disease.[23] Due to the “cytokine storm” also observed in poor outcome group, 19 (95.0%) of these patients were given glucocorticoid therapy. Wu et al previously found that the administration of methylprednisolone may have reduced the risk of death in patients with ARDS.[7] To our surprise, most of the severe patients treated with Traditional Chinese medicine (TCM) were eventually converted to good outcome, indicated this important effort on COVID-19. A large of clinical practice results indicated that TCM shows significant role in the patients with COVID-19. For the severe patients in the treatment of TCM, the mean length in hospital and the time of nucleic acid turning negative has been shortened by more than 2 days.[24] Yao et al. and Lu et al. analyzed the effect of Lian Hua Qing Wen Capsule in treatment of COVID- 19 patients and they found that this TCM could markedly relieve fever and cough and can promote the recovery.[25] Besides, the comprehensive evaluation and scientific research should be carried out on the effect of TCM on COVID-19.

Meantime, the risk factors related to the poor outcome included uncontrolled inflammation responses, infection, liver and kidney, cardiac dysfunction and hypoxia. The pathogenesis of COVID-19 is still studying. Cytokine storm and uncontrolled inflammation responses are thought to play important roles in the outcome of COVID-19.[26-30] External stimuli resulted to excessive immune response and the pathogenesis of the cytokine storm is complex which leaded to rapid disease progresses and high mortality. The inflammatory cytokine storm is closely correlated to the development and progression of ARDS.[31] Neutrophils play important role in chemokines and cytokines.[32] In our study, poor outcome had significantly higher neutrophil counts than good outcome group, which may leading to the cytokine storm. In addition, CD8+ T cells were significantly lower in poor outcome group. These results mean the important roles of CD8 T cells in COVID-19. Studies had shown that T-cell could inhibit the over-activation of innate immunity.[33] T cells can help to clear the SARS-CoV, and low T-cell response can result in pathological changes in mice with SARS-CoV.[34] The relevant mechanisms need to be further studied.

This study has some limitations. First, owing to the limited number of cases, only 114 severe patients were included. Second, this study was a single-center research and a larger cohort study of severe patients with SARS-CoV-2 from other cities in China and other countries would help to further describe the clinical characteristics and predict the risk factors about this disease.

In summary, we firstly reported this single-centered, prospective, observational study for short-term in severe patients with COVID-19. We found from univariate and multivariate Cox model that cytokine storm and uncontrolled inflammation responses, liver, kidney, cardiac dysfunction may play important roles in final outcome of severe ill patients with COVID-19. This will help clinicians to diagnose and treat severe patients.

## Data Availability

All data generated or analyzed during this study are available from corresponding author

## Author contributions

Cao Yang and Wei Yang designed the study and had full access to all data in the study and take responsibility for the integrity of data and the accuracy of the data analysis. Xiaobo Feng and Liang Ma contributed to data collection, literature searches, and writing of the manuscript. Xiaobo Feng and Peiyun Li had roles in data analysis and data interpretation. All authors contributed to data acquisition, clinical management, and reviewed and approved the final version of the manuscript. Xiaobo Feng, Peiyun Li and Liang Ma share first authorship and Cao Yang and Wei Yang are co-corresponding authors; the order in which they are listed was determined by workload.

## Declaration of interests

We declare no competing interests.

## Acknowledgments

We thank all the patients and their families involved in this study, as well as numerous civilians working together to fight against the SARS-CoV-2.

## Notes

### Competing Interest Statement

The authors have declared no competing interest.

### Funding Statement

None

